# When does rebleeding occur in subarachnoid hemorrhage? Revisiting rebleeding cases with a focus on blood pressure

**DOI:** 10.1101/2025.05.23.25328264

**Authors:** Hitoshi Kobata, Adan Tucker, Gemmalyn Sarapuddin, Makiko Kawakami

**Author notes:** Corresponding author: Hitoshi Kobata, Department of Neurosurgery, Tane General Hospital.

## Abstract

**BACKGROUND:** Early brain injury attributable to initial or recurrent bleeding is the leading cause of poor outcomes in patients with subarachnoid hemorrhage (SAH). This study investigated rebleeding immediately after SAH ictus, focusing on its timing and associated blood pressure (BP).

**METHODS:** Consecutive patients with spontaneous SAH treated from January 1999 to June 2022 were reviewed. Rebleeding was defined as a sudden decline in consciousness to a coma, sudden pupillary dilation with elevated BP, or increased SAH on head computed tomography (CT). The rebleeding timing was examined during each management phase. Demographic, radiological, and initial evaluation data were assessed for rebleeding and outcomes.

**RESULTS:** Among 940 patients (64% women, mean age 63.6 ± 13.2, 73% with a World Federation of Neurological Societies grade ≥4), rebleeding occurred in 221 cases (23.5%); 139 episodes in 121 patients (13.7%) before and 134 episodes in 112 patients after hospitalization (11.9%), and 19 patients (2.0%) in both. Rebleeding occurred more frequently in patients with worse neurological state, higher CT grades, and earlier arrivals. Systolic BP (SBP) was higher in patients with rebleeding (178 mmHg, IQR 140–204 mmHg) than in those without rebleeding (148 mmHg, IQR 100–180 mmHg) (*P*<0.001). Higher SBP was associated with increased rebleeding (OR 9.843; *P*<0.0001) and lower mortality (OR 0.0281; *P*=0.0084) but not with favorable outcomes (OR 1.686; *P*=0.22). When comparing the groups divided into 20 mmHg increments, the incidence of rebleeding, unfavorable outcomes, and mortality increased in the groups with SBP ≥181 mmHg, SBP ≤100 mmHg and >160 mmHg, and SBP ≤100 mmHg, respectively.

**CONCLUSIONS:** Rebleeding occurred in 23.5% of patients with SAH in the hyperacute phase, primarily before hospital arrival. Higher SBP was associated with rebleeding, and SBP of 101–160 mmHg was associated with favorable outcomes.

## INTRODUCTION

Preventing rebleeding with early resuscitation is essential for managing spontaneous subarachnoid hemorrhage (SAH). Rebleeding is a major determinant of outcomes, yet it could be preventable. Studies have focused on the importance and risk factors of rebleeding before securing the culprit lesion.^1–5^

Decades ago, before early surgery became widely adopted, the peak incidence of rebleeding was believed to occur between days 5 and 11 after onset.^6^ Subsequently, the International Cooperative Aneurysm Study identified that the highest rebleeding rate occurred within the first 24 h after initial SAH, reaching approximately 4.1%.^7^ Later studies found that rebleeding was the most frequent within 6 h^2,8^ or even within 2 h^4^ after ictus, leading some authors to recommend withholding angiography and aneurysm surgery during the first 6 h.^3,8^

In contrast, other researchers have argued that SAH should be treated with immediate surgery owing to its high incidence of early rebleeding.^9,10^ Protocol changes emphasizing emergency surgery have been associated with reduced rebleeding rates and improved outcomes.^11^ However, implementing a 24-h ultra-early aneurysm repair protocol would result in only a marginal reduction in rebleeding incidence.^12^

Nowadays, the primary cause of unfavorable outcomes in SAH is early brain injury (EBI) developing within 72 h after ictus.^13^ Advances in emergency medical systems and neurocritical care have enabled more critically ill patients with SAH to receive aggressive treatment, highlighting the increasing importance of managing SAH in the hyperacute phase. As up to one-fourth of patients with SAH die before arriving at the hospital,^14^ prehospital studies would be valuable. Although rebleeding before hospitalization or during transportation has been reported,^3–5,15–17^ comprehensive data on rebleeding throughout the entire course from ictus to securing the bleeding source remain limited. This study aimed to investigate rebleeding in the hyperacute phase of SAH immediately after ictus, with a particular focus on its timing and associated blood pressure (BP).

## METHODS

This single-center, retrospective, observational study was conducted at a tertiary critical care center, with approval by the Institutional Review Board of our institution (#2011-01), which waived the requirement for informed consent.

Prospectively registered all consecutive patients with spontaneous SAH from January 1999 to June 2022 were reviewed. Patients with aneurysms, dissecting lesions, or undetected bleeding sources, including SAH-induced cardiac arrest, were eligible. The database includes detailed information on ictus and transportation obtained from witnesses and paramedics.

SAH onset was defined as severe sudden headache, loss of consciousness, or both. Sentinel bleeding was not considered the onset. Rebleeding was defined as (i) a sudden decline in consciousness to a coma (Glasgow Coma Scale [GCS] score of ≤6), (ii) sudden pupillary dilation with elevated BP in comatose patients, or (iii) increased SAH amount on head computed tomography (CT) when a prior CT scan was available for comparison.

The timing of rebleeding was classified as before the contact of emergency medical services (EMS) at the site (pre-EMS), during ambulance transport (ambulance), in the emergency department (ED), initial CT imaging (CT), CT angiography (CTA), diagnostic digital cerebral angiography (DSA), in the operating room before craniotomy (OP), and during observation without emergency treatment of the bleeding source (others).

Along with prompt neurological examination, comatose patients with suspected SAH underwent endotracheal intubation in the ED under deep sedation and analgesia, with systolic BP (SBP) targeted at ≤120 mmHg under continuous monitoring of radial artery pressure. Midazolam, fentanyl, buprenorphine, vecuronium, or rocuronium were initially administered, and propofol or thiamylal was added if necessary. Nicardipine was continuously infused intravenously if these measures did not reduce the SBP to the target level. Good-grade patients (GCS score of ≥13) underwent the same management after informed consent was provided

Etilefrine hydrochloride was administered if the SBP decreased excessively below 100 mmHg, and vasopressors were continuously infused when required. Cardiac function was assessed simultaneously in the ED using electrocardiography and echocardiography.

Patients underwent head CT, CTA, or DSA after stabilizing vital signs. Since 2006, diagnostic DSA has generally not been performed if CTA identified the culprit lesion. Patients with cardiac arrest upon arrival underwent head CT to determine the etiology. Successfully resuscitated patients with SAH underwent CTA or DSA.

In principle, radical treatment was immediately performed to secure the culprit lesions when indicated, regardless of the time or day of the week, primarily via surgical clipping. Good-grade patients arriving at midnight may undergo radical treatment the following morning. Tranexamic acid or external ventricular drainage was not indicated before securing the bleeding source.

The following data were collected: age, sex, presence or absence of witnesses, onset-arrival and -treatment time, GCS score and modified World Federation of Neurological Societies (mWFNS) grade (Supplemental Table 1)^18^ upon arrival, pupillary findings, SBP, diastolic BP (DBP), heart rate, blood glucose (BS) and serum potassium (K) levels, plasma D-dimer level, location and shape of the bleeding source, admission route (directly from the site or inter-hospital transport), clinical signs indicating rebleeding events, and SBP immediately before endotracheal intubation if recorded.

Outcomes were assessed using the modified Rankin Scale (mRS) at 6 months and dichotomized into favorable (scores 0–2) or unfavorable (scores 3–6), which was obtained from records from our hospital or the rehabilitation facilities where the patients were transferred.

### Statistical analyses

Baseline characteristics are summarized as numbers (%) for categorical variables and compared using the chi-square test or Fisher’s exact test, as appropriate. Continuous variables are summarized as means with standard deviations or medians with interquartile ranges (IQRs) based on the normality of the data. Patients with SBP between 101 and 200 mmHg were categorized into 20 mmHg increments and evaluated for association with rebleeding and outcomes, together with those with SBP ≤100 and ≥201 mmHg.

The Wilcoxon/Kruskal–Wallis test was used for between-group comparisons. Excluding confounding factors, clinical variables in the univariate analysis (*P*<0.25) were selected and entered into a multivariate binary regression model to ascertain independent predictors of rebleeding. Two-sided *P*<0.05 was considered statistically significant. All analyses were performed using JMP Pro, version 17.0.0 (SAS Institute Inc., Cary, NC, USA).

## RESULTS

Among 940 patients (mean age 63.6 ± 13.2 years, 64% women, 73% with a mWFNS grade ≥ 4), rebleeding occurred in 221 patients (23.5%) (Table 1). Patients with rebleeding exhibited worse mWFNS grades, worse modified Fisher (mFisher) CT grades, higher SBP and DBP, higher BS and lower serum K levels, higher BS/K ratio, dissecting lesions, shorter onset-arrival and -treatment times, and a trend toward bilaterally dilated pupils and higher plasma D-dimer levels than those without rebleeding. Patients without rebleeding underwent radical treatment less frequently but had more favorable outcomes than those with rebleeding.

**TABLE 1.** Demographic and clinical information in 940 patients with SAH.

| Variable | Rebleeding<br>(n=221) | No rebleeding<br>(n=719) | *P* value |
| --- | --- | --- | --- |
| Age in yrs, mean ± SD | 63.3 ± 12.5 | 63.7 ± 13.4 | 0.71 |
| Female sex | 133 (60.2) | 469 (65.2) | 0.17 |
| modified WFNS grade |  |  | <0.0001 |
| I (GCS 15) | 7 (4.9) | 136 (95.1) |  |
| II (GCS 14) | 10 (13.3) | 65 (86.7) |  |
| III (GCS 13) | 7 (18.9) | 30 (81.1) |  |
| IV (GCS 7-12) | 33 (23.4) | 108 (76.6) |  |
| V (GCS 3-6) | 149 (42.6) | 201 (57.4) |  |
| Cardiac arrest | 15 (7.7) | 179 (92.3) |  |
| Lack of light rection | 99 (44.8) | 311 (43.3) | 0.69 |
| Bilateral pupillary dilatation | 183 (25.5) | 40 (18.1) | 0.074 |
| modified Fisher grade |  |  | <0.0001 |
| 0 | 3 (75.0) | 1 (25.0) |  |
| 1 | 15 (10.9) | 123 (89.1) |  |
| 2 | 21 (31.3) | 46 (68.7) |  |
| 3 | 129 (23.4) | 428 (76.8) |  |
| 4 | 53 (30.5) | 121 (69.5) |  |
| Systolic BP (mmHg) | 178 (140–204) | 148 (100–180) | <0.0001 |
| Systolic BP (Excl cardiac arrest) | 180 (150–209.5) | 161.5 (136–190) | <0.0001 |
| Diastolic BP (mmHg) | 90 (78–110) | 82 (54–100) | <0.0001 |
| Heart rate (bpm) | 78 (66–98) | 80 (60–96) | 0.19 |
| Blood glucose (ng/dL) | 200 (173–229) | 187 (151–239) | 0.0092 |
| Serum K (mEq/L) | 3.3 (3.0–3.6) | 3.4 (3.1–3.8) | <0.0001 |
| Blood glucose/serum K | 61.1 (51.5–72.9) | 53.2 (42.4–69.3) | <0.0001 |
| Plasma D-dimer (μg/mL) * | 4.5 (1.9–11.0) | 3.5 (1.3–11.9) | 0.098 |
| Neurogenic stand myocardium | 20 (25.0) | 201 (23.4) | 0.74 |
| Location of the lesion |  |  | 0.42 |
| Anterior | 171 (80.3) | 484 (82.7) |  |
| Posterior | 42 (29.4) | 101 (17.3) |  |
| Shape of the lesion |  |  | 0.0064 |
| Saccular aneurysm | 185 (86.9) | 544 (93.0) |  |
| Dissecting lesion | 28 (13.1) | 41 (7.0) |  |
| Onset-arrival time (min) † | 41 (29–73) | 44 (31–104) | 0.021 |
| Onset-treatment time (min) † | 165 (120–230) | 205 (150–312.5) | <0.0001 |
| Interhospital transfer | 46 (21.5) | 156 (21.6) | 0.99 |
| Treatment |  |  | 0.0004 |
| Clipping | 149 (67.2) | 407 (56.8) |  |
| IVR | 21 (9.5) | 44 (6.2) |  |
| None | 51 (23.1) | 265 (37.0) |  |
| Favorable outcomes (mRS 0–2) ‡ | 51 (27.6) | 283 (39.5) | 0.0013 |
| Mortality | 90 (40.7) | 274 (38.3) | 0.51 |

Data are expressed as the absolute number (%) of patients or the median with interquartile range (IQR) unless otherwise stated.
\* Data available from 753 patients.
† Onset time was identified in 721 patients.
‡ Outcome was assessed in 937 patients excluding 3 transferred to another hospital without treatment for the culprit lesion.
BP indicates blood pressure; GCS, Glasgow Coma Scale; IVR, interventional radiology; mRS, modified Rankin Scale; WFNS, World Federation of Neurological Society.

Fig. 1 shows the number of patients with favorable or unfavorable outcomes according to the rebleeding timing. Rebleeding occurred 273 times in 221 patients—once in 181, twice in 30, three times in 3, and four times in 2 patients. Rebleeding occurred 139 times in 129 patients (13.7%) before hospitalization and 134 times in 112 patients (11.9%) after hospitalization (Table 2). Nineteen patients (2.0%) rebled both before and after hospitalization. Thirty-seven patients rebled during inter-hospital transport after a CT diagnosis of SAH. Ten of the 20 patients in the others rebled while being withheld from aggressive treatment because of poor clinical conditions.

**Figure 1.**
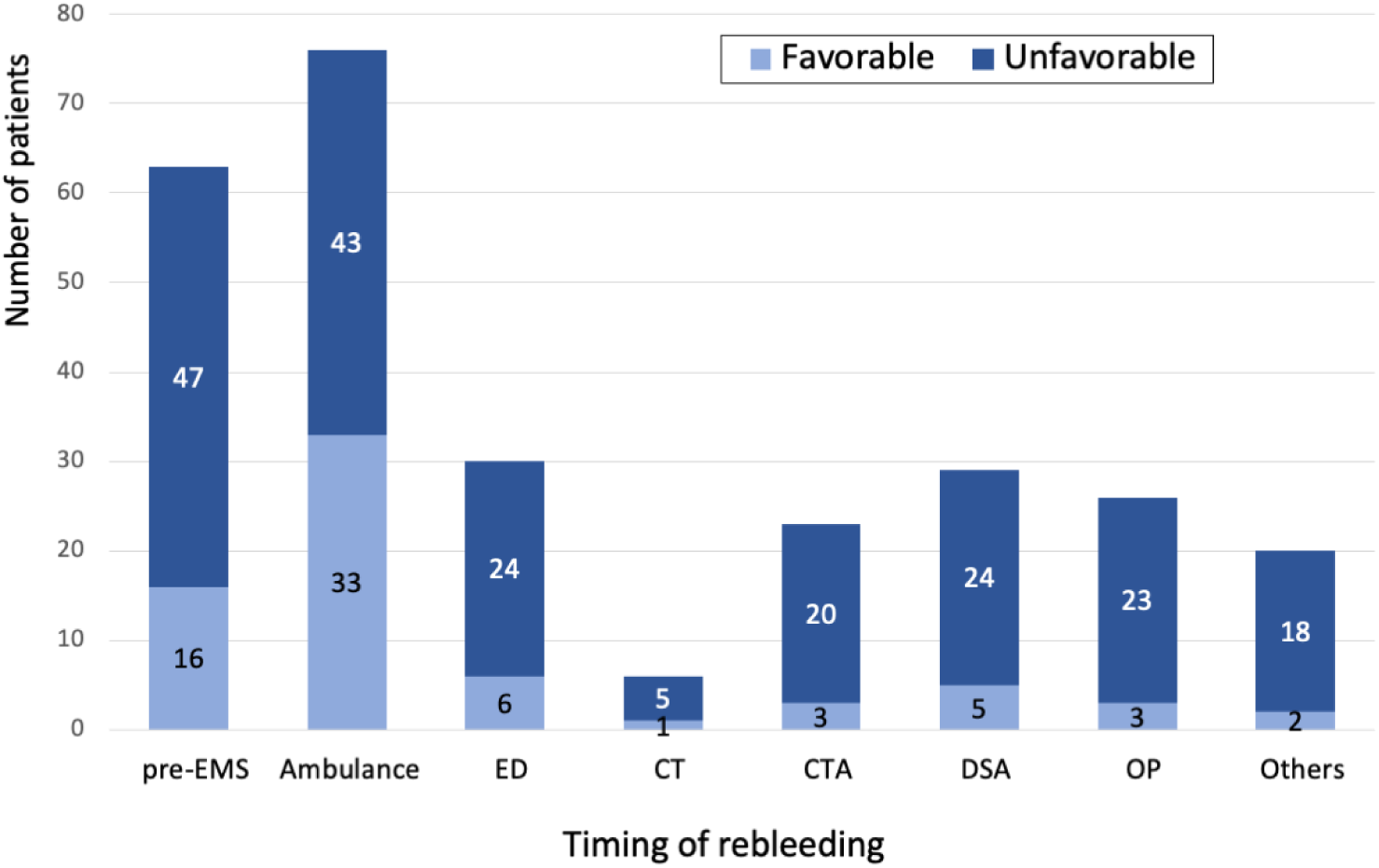
Number of patients according to outcome in each rebleeding phase. Rebleeding was most common in ambulance1, followed by pre-EMS, accounting for over half of the patients. Most patients had unfavorable outcomes, regardless of the rebleeding phase.

**TABLE 2.** Timing of rebleeding and outcomes in 221 patients compared with patients without rebleeding.

| Timing of rebleeding | Incidence of rebleeding | Outcomes |  |  |  |  |  |
| --- | --- | --- | --- | --- | --- | --- | --- |
|  |  | Unfavorable |  | P value | Mortality |  | P value |
|  |  | Rebleeding | No rebleeding |  | Rebleeding | No rebleeding |  |
| Overall rebleeding | 273 (100) | 160 (72.4) | 433 (60.5) | 0.0013 | 90 (40.7) | 274 (38.3) | 0.51 |
| Before arrival | 139 (50.9) | 83 (63.3) | 510 (63.1) | 0.79 | 39 (30.2) | 325 (40.2) | 0.031 |
| Pre-EMS | 63 (23.1) | 47 (74.6) | 546 (62.5) | 0.054 | 22 (34.9) | 342 (39.1) | 0.51 |
| Ambulance | 76 (27.8) | 43 (56.6) | 560 (63.9) | 0.21 | 20 (26.3) | 344 (40.0) | 0.019 |
| After arrival | 134 (49.1) | 92 (82.1) | 501 (60.7) | <0.0001 | 65 (58.0) | 299 (36.2) | <0.0001 |
| ED | 30 (11.0) | 24 (80.0) | 569 (62.7) | 0.056 | 13 (43.3) | 351 (38.7) | 0.61 |
| CT | 6 (2.2) | 5 (83.3) | 588 (63.2) | 0.42 | 3 (50.0) | 361 (38.8) | 0.68 |
| CTA | 23 (8.4) | 20 (87.0) | 573 (62.7) | 0.016 | 18 (78.3) | 346 (37.9) | <0.0001 |
| DSA | 29 (10.6) | 24 (82.8) | 569 (62.7) | 0.030 | 19 (65.5) | 345 (38.0) | 0.0028 |
| OP | 26 (9.5) | 23 (88.5) | 570 (62.6) | 0.0064 | 13 (50.0) | 351 (38.5) | 0.24 |
| Others | 20 (7.3) | 18 (90.0) | 575 (62.7) | 0.01 | 17 (85.0) | 347 (37.8) | <0.0001 |
| Multiple | 40 (14.7) | 35 (87.5) | 558 (62.2) | 0.0007 | 27 (67.5) | 337 (37.6) | 0.0001 |

Data are expressed as the absolute number of patients (% of all 940 patients) or episodes (% of 273 rebleeding).
CT indicates computed tomography; CTA, computed tomography angiography; DSA, digital subtraction angiography; ED, emergency department; EMS, emergency medical service; OP, operation.

Overall, rebleeding was associated with unfavorable outcomes but not mortality. Rebleeding before hospitalization exhibited no association with unfavorable outcomes but was linked to lower mortality. Specifically, pre-EMS rebleeding was associated with a trend toward unfavorable outcomes, whereas rebleeding during ambulance transport was associated with lower mortality. In-hospital rebleeding was associated with both unfavorable outcomes and higher mortality. Rebleeding in the ED showed a trend toward unfavorable outcomes but not for mortality. Rebleeding during CTA, DSA, and others was associated with unfavorable outcomes and mortality, and during OP was associated with unfavorable outcomes but not with mortality. Multiple rebleeding events were associated with both unfavorable outcomes and mortality.

Rebleeding occurred more frequently as the mWFNS scores worsened, regardless of whether before or after hospital arrival (Fig. 2). Several patients successfully resuscitated after cardiac arrest also rebleed.

**Figure 2.**
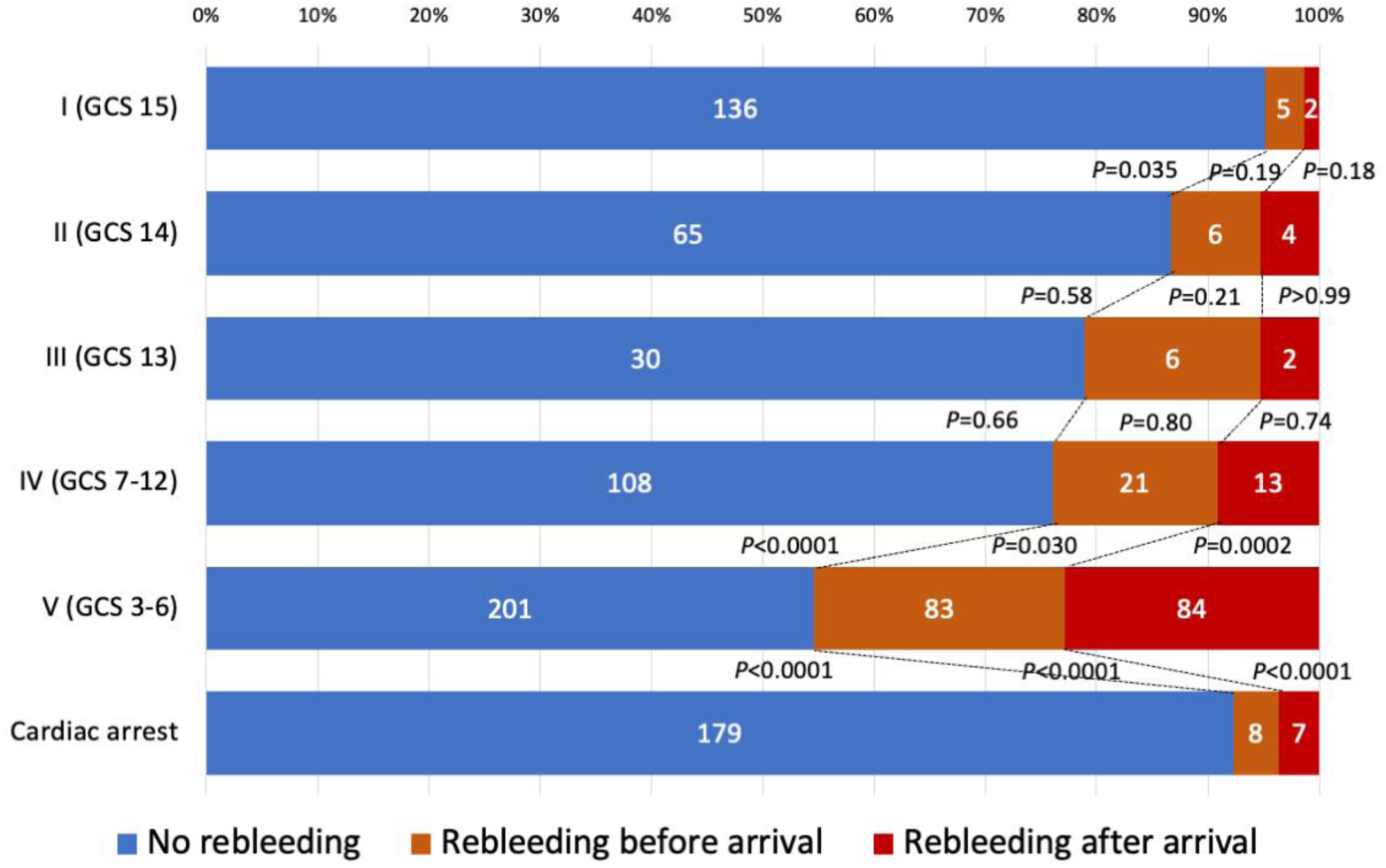
Neurological grade upon arrival and rebleeding. The frequency of rebleeding increased as the mWFNS score worsened, except in cardiac arrest. Rebleeding occurred in 4.9%, 13.3%, 18.9%, 23.4%, 42.6%, and 7.7% of patients with mWFNS grades I, II, III, IV, V, and cardiac arrest, respectively. The difference in overall rebleeding was significant between mWFNS grades I and II (*P*=0.035) and IV and V (*P*<0.0001). Rebleeding before and hospital arrival was significantly more frequent in patients with mWFNS grade V than in those with grade IV (*P*=0.030 and 0.0002, respectively). Patients successfully resuscitated from cardiac arrest also rebled less frequently compared with grade V, regardless of the timing (all *P*<0.0001).

Onset-arrival time was 43 min (IQR 30−94.8 min) for 721 patients with identified onset time. Patients with rebleeding arrived significantly earlier than in those without rebleeding. The time was significantly shorter in patients with rebleeding after arrival than those without. However, no difference in time was observed in patients with prehospital rebleeding (Fig. 3). SBP was higher in patients with overall rebleeding than in those without, regardless of rebleeding before or after arrival (Fig. 4).

**Figure 3.**
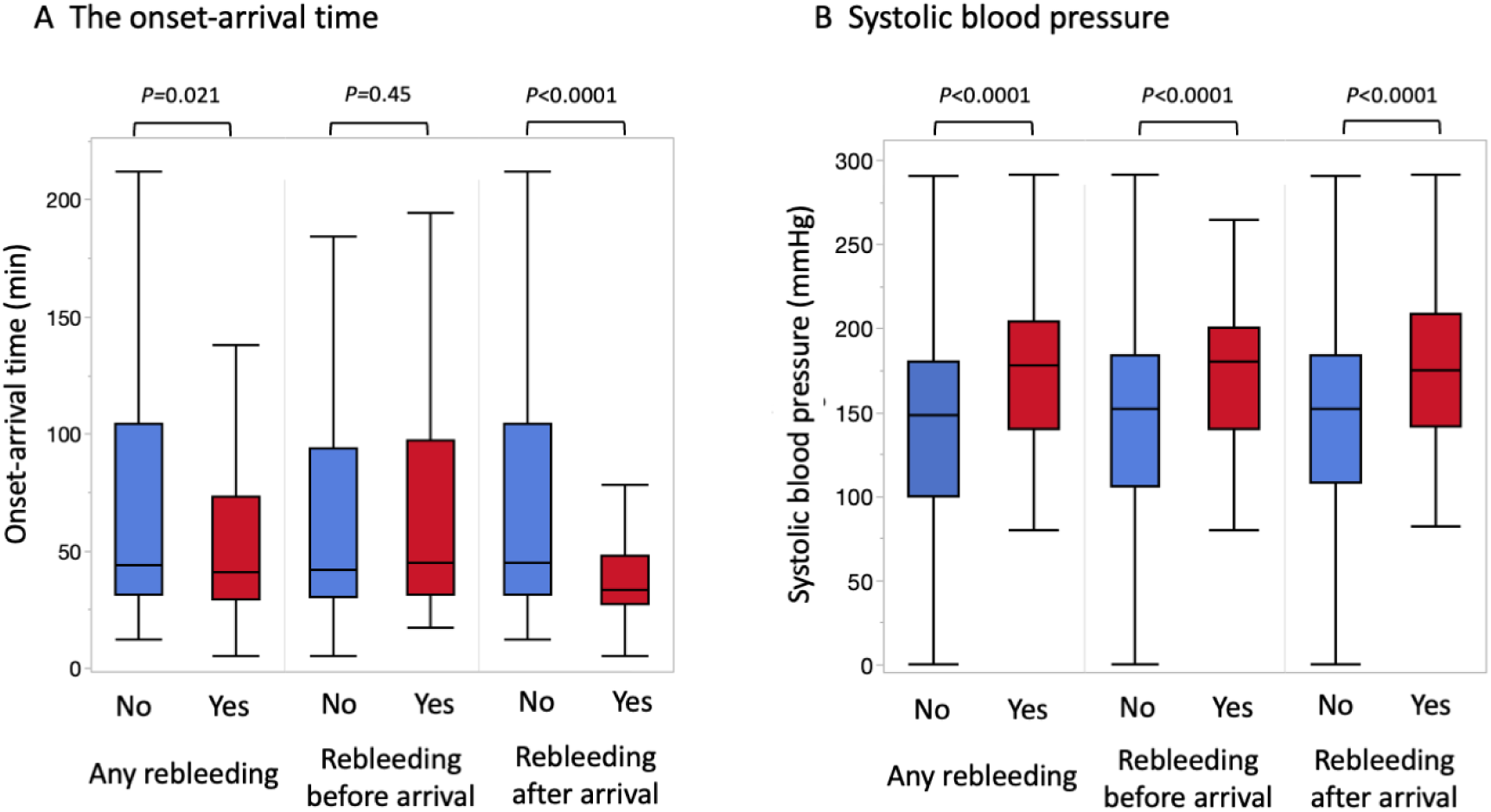
Box plot showing the relationship between onset-arrival time, systolic blood pressure, and rebleeding. **A,** The onset-arrival time was significantly shorter in patients with rebleeding than in those without rebleeding (41 min, IQR 29*–*73 min vs. 44 min, IQR 31*–*104 min; *P*=0.021). The time was not significantly different between patients with rebleeding before arrival and those without (45 min, IQR 31*–*97 min vs. 42 min, IQR 30*–*94 min; *P*=0.45), but significantly shorter in patients with rebleeding after arrival than in those without (33 min, IQR 27*–*48 min vs. 45 min, IQR 34*–*104 min; *P*<0.0001). **B,** Systolic blood pressure (SBP) was higher in patients with overall rebleeding (178 mmHg, IQR 140*–*204 mmHg vs. 148 mmHg, IQR 100–180 mmHg; *P*<0.001), in those with rebleeding before arrival (180 mmHg, IQR 140.5*–*200 mmHg vs. 152 mmHg, IQR 106*–*184 mmHg; *P*<0.0001), and after arrival (175 mmHg, IQR 141.5*–*208.5 mmHg vs. 152 mmHg, IQR 108*–*184 mmHg; *P*<0.0001) than in those without rebleeding.

**Figure 4.**
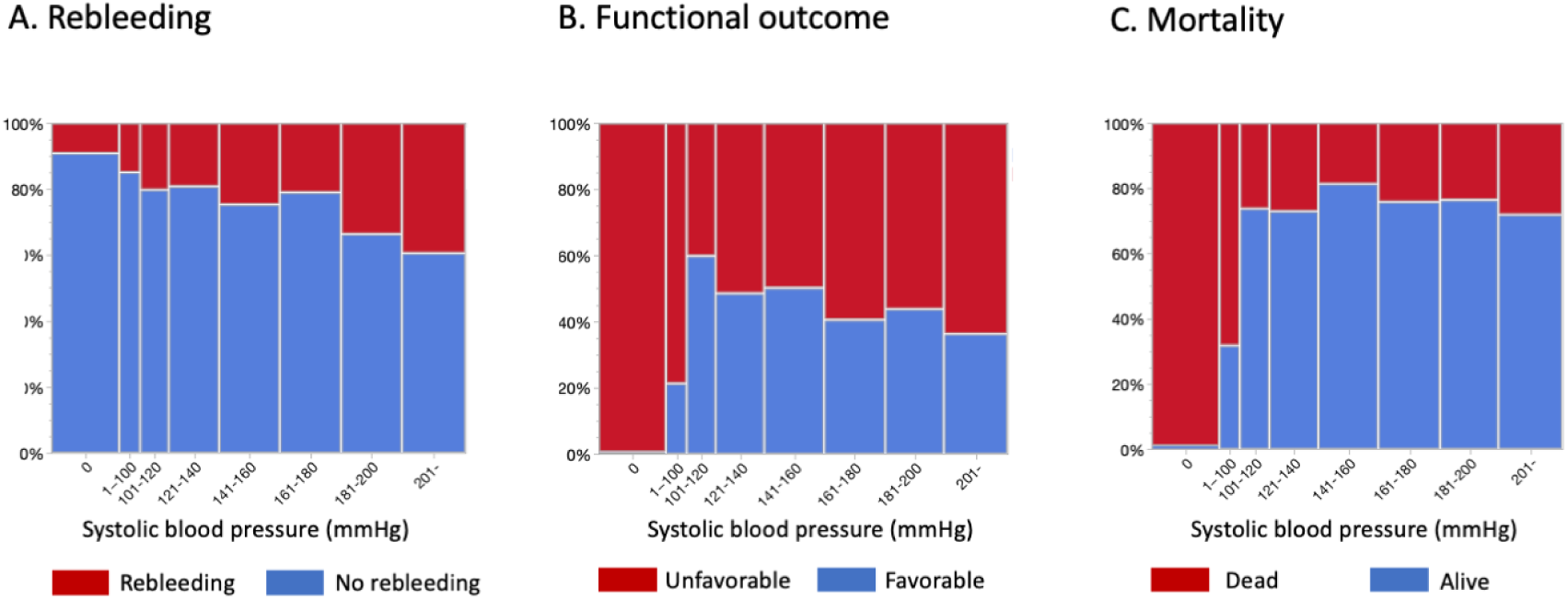
Association of systolic blood pressure with rebleeding, functional outcome, and mortality A, Higher systolic blood pressure (SBP) was associated with an increased rebleeding (*P*<0.0001). **B,** Patients with an SBP of 101–120 mmHg exhibited the highest rate of favorable outcomes. SBP ≤100 mmHg and SBP ≥161 mmHg were associated with unfavorable outcomes. **C,** Mortality was associated with SBP ≤100 mmHg, but not with higher SBP. See Table 4 for odds ratios and 95% confidence intervals.

A worse mWFNS grade was associated with higher SBP; the difference was significant between grades I and II, and IV and V (Supplemental Fig. 1). Generally, patients with rebleeding underwent more aggressive treatment for the culprit lesion, particularly those with prehospital rebleeding, than those without. Patients with rebleeding during CTA and others were less likely to undergo definitive treatment (Supplemental Table 2).

Patients with rebleeding presented with more severe hemorrhage, as indicated by the mFisher CT grade; significantly higher incidence of grade 4 and lower incidence in combined grades 1 and 2 compared to those without rebleeding. (Supplemental Fig. 2).

Higher SBP was associated with increased rebleeding (odds ratio [OR] 10.829; *P*<0.0001), favorable outcomes (OR 0.0869; *P*<0.001), and lower mortality (OR 0.00711; *P*<0.001) (Supplemental Figs. 3A-C). When patients with cardiac arrest were excluded, a higher SBP was still associated with increased rebleeding (OR 9.843; *P*<0.0001), unrelated to favorable outcomes (OR 1.686; P< 0.22), but with lower mortality (OR 0.0281; *P*=0.0084) (Supplemental Figs. 3D-F).

The higher SBP groups were associated with increased rebleeding when classified into zero, 1–100, 101–120, 121–140, 141–160, 161–180, 181–200, and ≥201 mmHg groups (Fig. 2C). The 101–120 mmHg SBP group exhibited the highest rate of favorable outcomes (Fig. 2D). The mortality was higher in groups with SBP ≤100 mmHg but did not differ significantly between groups with SBP ≥101 mmHg (Fig. 2E).

Table 3 shows the relationship between SBP and rebleeding, functional outcome, and mortality using the SBP 101–120 mmHg group as a reference. The rebleeding rate was lower in the zero mmHg SBP group and higher in the ≥181 mmHg SBP groups. Unfavorable outcomes significantly increased in the ≤100 and ≥161 mmHg SBP groups. The mortality rate was higher in the ≤100 mmHg SBP groups, with no significant differences between ≥101 mmHg SBP groups.

**TABLE 3.**
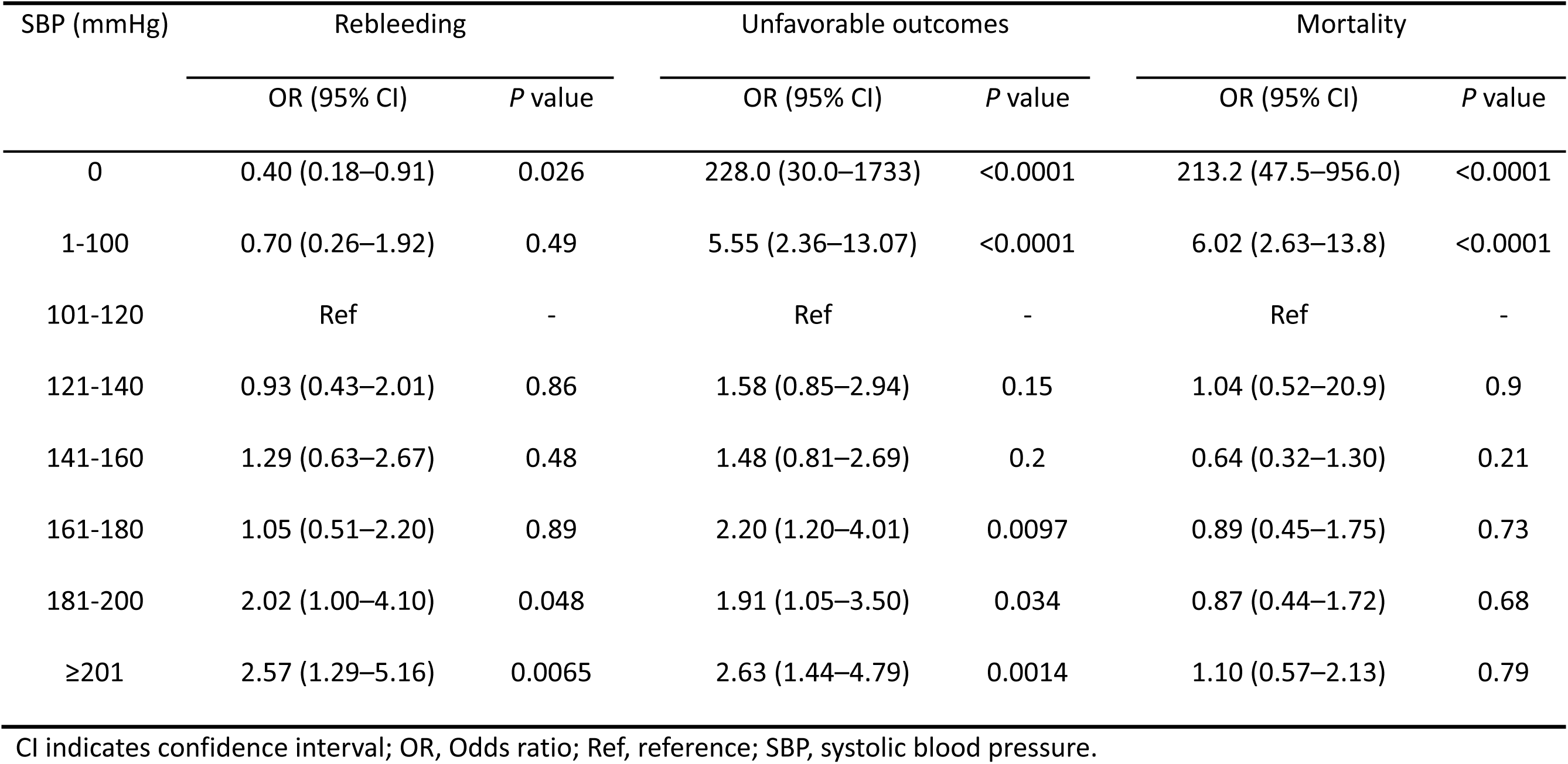
Relation of systolic blood pressure to rebleeding, functional outcome, and mortality.

The incidences of rebleeding, functional outcomes, and mortality are presented among cohorts dichotomized based on 100, 120, 140, 160, 180, and 200 mmHg SBP as thresholds (Table 4; Supplemental Figs. 4–6). In all groups with SBP thresholds of ≥120 mmHg, the rebleeding rate significantly increased in the higher SBP groups. Unfavorable outcomes significantly increased in the lower SBP group with thresholds of 100 mmHg and decreased in the lower SBP groups with thresholds of 160 and 200 mmHg. Mortality significantly increased in the lower SBP groups with thresholds of ≤140 mmHg.

**TABLE 4.** Incidence of rebleeding, functional outcome, and mortality among cohorts dichotomized using SBP 100, 120, 140, 160, 180, and 200 mmHg as thresholds.

| SBP | Rebleeding | *P* value | Unfavorable | *P* value | Mortality | *P* value |
| --- | --- | --- | --- | --- | --- | --- |
| ≤100 (n = 47) | 7 (14.9) | 0.085 | 37 (80.4) | 0.0006 | 32 (69.6) | <0.0001 |
| >100 (n = 727) | 197 (27.1) |  | 397 (54.8) |  | 176 (24.3) |  |
| ≤120 (n =112) | 20 (17.9) | 0.027 | 63 (56.3) | 0.99 | 49 (43.8) | <0.0001 |
| >120 (n = 662) | 184 (27.8) |  | 371 (56.3) |  | 159 (24.1) |  |
| ≤140 (n = 223) | 41 (18.4) | 0.0014 | 120 (53.8) | 0.38 | 79 (35.4) | 0.0007 |
| >140 (n = 551) | 163 (29.6) |  | 314 (57.3) |  | 129 (23.5) |  |
| ≤160 (n =358) | 74 (20.7) | 0.0009 | 187 (52.2) | 0.035 | 104 (29.1) | 0.23 |
| >160 (n = 416) | 130 (31.3) |  | 247 (59.8) |  | 104 (25.2) |  |
| ≤180 (n =498) | 103 (20.7) | <0.0001 | 269 (54.1) | 0.103 | 138 (27.8) | 0.51 |
| >180 (n = 276) | 101 (36.6) |  | 165 (60.2) |  | 70 (25.6) |  |
| ≤200 (n = 631) | 148 (23.5) |  | 343 (54.6) |  | 168 (26.8) |  |
|  |  | 0.0001 |  | 0.05 |  | 0.77 |
| >200 (n = 143) | 56 (39.2) |  | 91 (63.6) |  | 40 (28.0) |  |

Data are expressed as the absolute number (%) of patients.
SBP indicates systolic blood pressure.

Among the 624 patients whose SBP was recorded immediately before endotracheal intubation, the median SBP was 117 mmHg (IQR 100–134 mmHg). However, SBP of ≤120 and ≤140 mmHg could not be achieved in 257 (41.1%) and 115 (18.4%) patients, respectively. Of the 11 patients with SBP remaining ≥201 mmHg, 6 (58%) rebled after arrival, and 2 rebled both before and after hospitalization. Similarly, of the 22 patients with SBP of ≥181 mmHg, 10 (40%) rebled, 8 after and 5 before hospitalization, and 3 both before and after hospitalization.

In the multivariate analysis, rebleeding was associated with the worse mWFNS grade (*P*<0.0001) and dissecting lesions (*P*=0.0041). Heart rate (*P*=0.053) and mFisher CT grade (*P*=0.073) showed a trend toward an association with rebleeding (Supplemental Table 3).

## DISCUSSION

Our study revealed that rebleeding occurred in 23.5% of patients with SAH in the hyperacute phase, primarily in the prehospital phase. Rebleeding was more frequent within a shorter time after ictus and increased proportionally with SAH severity. Higher SBP was significantly associated with an increased risk of rebleeding. Patients with an SBP of 101–160 mmHg were more likely to relate favorable outcomes, whereas mortality increased at SBP ≤100 mmHg and did not differ at SBP ≥101 mmHg. These findings suggest that an SBP of 101–160 mmHg is a reasonable target before securing the culprit lesion. Shorter onset-arrival time, worse neurological grades and related findings, worse CT grades, higher SBP and DBP, and dissecting lesions were associated with rebleeding. Bilaterally dilated pupils and elevated plasma D-dimer levels showed a trend toward rebleeding. Multivariate analysis identified the mWFNS grade and dissecting lesions as significant factors.

Rebleeding is associated with mortality and morbidity in patients with SAH.^1,3,5,19^ Rebleeding rates during the hyperacute phase vary across study cohorts, ranging from 4.9%^5^ to 22.0%.^20^ The reported incidence of rebleeding is 4.3%^8^ to 18.4%^3^ before admission and 5.9%^10^ to 16.3%^15^ after admission. The overall rebleeding rate of 23.5% in the current study was high, primarily because of the inclusion of prehospital rebleeding and a high volume of poor-grade SAH immediately after ictus.

A recent meta-analysis confirmed the association between rebleeding and worse neurological grades, as assessed using the Hunt and Hess or WFNS grades.^21,22^ Risk factors for pretreatment rebleeding included SBP of >160 mmHg, seizure or loss of consciousness at ictus, sentinel headache, Fisher or mFisher CT grade, associated intraventricular or intracerebral hemorrhage, subdural hematoma, aneurysm size of >10 mm,^21^ aneurysm location, and hydrocephalus.^23^ Others reported dissecting lesions,^24^ premorbid hypertension,^25^ age, multiple aneurysms, heart disease,^5^ and mean BP.^26^ Additionally, pupillary dilation and high plasma D-dimer levels are linked to SAH severity.^27,28^ A systematic review of prediction models for pretreatment rebleeding following SAH concluded that no models are currently recommended for clinical use.^23^

Findings from the present study align with recent meta-analyses while also offering novel insights. Most rebleeding events occurred prehospital, immediately after ictus. High SBP in the hyperacute phase appeared to be a consequence of, and a risk factor for, rebleeding.

This study revealed lower mortality rates in patients with prehospital rebleeding. This paradox may be attributed to a selection bias favoring radical treatment. Patients with prehospital rebleeding were more likely to undergo definitive treatments expecting good recovery through ultra-early intervention. Although this aggressive approach was lifesaving, it did not improve neurological outcomes. Extravascular leakage of contrast medium during CTA or DSA may exacerbate damage and discourage further treatment.^2,3,29^

Hypertension could be a modifiable risk factor for rebleeding. Observational studies have demonstrated an association between elevated BP and rebleeding, particularly at SBP of >160 mmHg.^4,15,30^ A meta-analysis indicated that SBP of >160 mmHg was more closely associated with rebleeding than SBP of >140 mmHg.^31^ Previous guidelines suggested a target SBP of <160 mmHg^32^ or <180 mmHg.^33^ However, current guidelines do not recommend a specific target SBP before aneurysm treatment because of insufficient evidence.^34,35^ A meta-analysis examining SBP thresholds of 140, 160, 180, and 200 mmHg failed to determine a definitive association between elevated SBP and rebleeding risk during an unsecured period. Additionally, all reviewed studies exhibited a moderate-to-severe risk of bias.^19^

CT perfusion studies demonstrated that cerebral autoregulation deteriorates with the severity of EBI. Maintaining a physiological BP is essential for preserving cerebral blood flow, particularly in patients with poor-grade SAH.^36^ The target SBP should be determined by balancing the risk of rebleeding with sufficient cerebral circulation, considering premorbid SBP.

Strategies for BP reduction are essential. Vasodilators may increase intracranial pressure and decrease cerebral blood flow by inducing cerebral vasodilation. In our hospital, patients with poor-grade SAH were managed with deep sedation and strict BP control immediately after hospitalization, targeting an SBP of ≤120 mmHg, followed by prompt radical treatment.^37^ Immediate general anesthesia in the ED and emergent definitive treatment are feasible and contribute to a lower rebleeding rate (4.7%) in all patients with SAH^24^ and a significant reduction in rebleeding and mortality in good-grade SAH (WFNS grades I–III) compared with nicardipine-based management.^38^ Short-term BP reduction using neuroprotective drugs until securing the culprit lesion may be tolerable and beneficial. However, some patients in our study did not achieve the target SBP, even with aggressive management. Refractory hypertension was associated with a high rate of intermittent or ongoing rebleeding.

Whether the ultra-early securing strategy reduces rebleeding remains debatable. ^11,12^ Differences in SAH severity and the time from initial onset may account for discrepancies across studies. Our cohort was characterized by a high prevalence of poor-grade SAH, arriving exceptionally early after ictus. The results highlight the importance of prehospital rebleeding, especially during emergency transport. Intervention with EMS is desirable, at least for patients presenting with a typical SAH.

This study has some limitations and biases. First, this was a single-center retrospective study. Although patients were treated following a consistent treatment regimen throughout the study period, the initial management may differ depending on the attending physician. In addition, some data on the variables could not be collected. Second, rebleeding was primarily diagnosed based on clinical findings without a radiological study. Rebleeding may be underestimated in minor hemorrhage or overestimated in epilepsy. Third, the difficulties in treating the culprit lesion and delayed cerebral ischemia were not considered in the outcome assessment. Finally, the outcome assessment was not blinded.

## CONCLUSIONS

Rebleeding occurred in 23.5% of patients with SAH in the hyperacute phase, primarily before hospital arrival. High SBP was significantly associated with rebleeding, and favorable outcomes were more common in patients with an SBP of 101–160 mmHg. Although the target SBP should be determined individually, this study suggests that this SBP range is reasonable before securing the culprit lesion.

## Data Availability

N/A

## Non-standard Abbreviations and Acronyms

BP: blood pressure
CT: computed tomography
CTA: computed tomography angiography
DSA: digital cerebral angiography
EBI: early brain injury
ED: emergency department
EMS: emergency medical services
GCS: Glasgow Coma Scale
IQR: interquartile range
mWFNS: modified World Federation of Neurological Societies
OR: odds ratio
SAH: subarachnoid hemorrhage
SBP: systolic blood pressure
WFNS: World Federation of Neurological Societies

## Acknowledgments

The authors thank all paramedics and medical staff involved in this study.

## Supplemental Material

Tables S1-S3

Figures S1-S6

## REFERENCES

1. Broderick JP, Brott TG, Duldner JE, Tomsick T, Leach A: Initial and recurrent bleeding are the major causes of death following subarachnoid hemorrhage. Stroke 1994;25(7):1342–1347. doi: 10.1161/01.str.25.7.1342

2. Hillman J, von Essen C, Leszniewski W, Johansson I. Significance of “ultra-early” rebleeding in subarachnoid hemorrhage 1988; 68(6):J Neurosurg 901–917. doi: 10.3171/jns.1988.68.6.0901

3. Fujii Y, Takeuchi S, Sasaki O, Minakawa T, Koike T, Tanaka R: Ultra-early rebleeding in spontaneous subarachnoid hemorrhage. J Neurosurg 1996;84(1):35–42. doi: 10.3171/jns.1996.84.1.0035

4. Ohkuma H, Tsurutani H, Suzuki S: Incidence and significance of early aneurysmal rebleeding before neurosurgical or neurological management. Stroke 32:1176–1180, 2001. doi: 10.1161/01.str.32.5.1176

5. Horie N, Sato S, Kaminogo M, Morofuji Y, Izumo T, Anda T, Matsuo T. Impact of perioperative aneurysm rebleeding after subarachnoid hemorrhage. J Neurosurg. 2019;133(5):1401–1410. doi: 10.3171/2019.6.JNS19704

6. Locksley HB. Natural history of subarachnoid hemorrhage, intracranial aneurysms and arteriovenous malformations. Based on 6368 cases in the cooperative study. J Neurosurg. 19660;25(2):219–239. doi: 10.3171/jns.1966.25.2.0219

7. Kassell NF, Torner JC: Aneurysmal rebleeding: a preliminary report from the Cooperative Aneurysm Study. Neurosurgery1983;13(5):479–481

8. Inagawa T. Ultra-early rebleeding within six hours after aneurysmal rupture. Surg Neurol. 1994;42(2):130–134. doi: 10.1016/0090-3019(94)90373-5

9. Laidlaw JD, Siu KH. Ultra-early surgery for aneurysmal subarachnoid hemorrhage: outcomes for a consecutive series of 391 patients not selected by grade or age. J Neurosurg. 2002;97(2):250–258; discussion 247-249. doi: 10.3171/jns.2002.97.2.0250

10. van Donkelaar CE, Bakker NA, Veeger NJ, Uyttenboogaart M, Metzemaekers JD, Luijckx GJ, Groen RJ, van Dijk JM. Predictive factors for rebleeding after aneurysmal subarachnoid hemorrhage: Rebleeding Aneurysmal Subarachnoid Hemorrhage Study. Stroke 2015;46(8):2100–2106. doi: 10.1161/STROKEAHA.115.010037

11. Park J, Woo H, Kang DH, Kim YS, Kim MY, Shin IH, Kwak SG. Formal protocol for emergency treatment of ruptured intracranial aneurysms to reduce in-hospital rebleeding and improve clinical outcomes. Journal of neurosurgery J Neurosurg. 2015;122(2):383–391. doi:10.3171/2014.9

12. Linzey JR, Williamson C, Rajajee V, Sheehan K, Thompson BG, Pandey AS: Twenty-four-hour emergency intervention versus early intervention in aneurysmal subarachnoid hemorrhage. J Neurosurg 2018;128(5):1297–1303. doi: 10.3171/2017.2.JNS163017

13. Lauzier DC, Jayaraman K, Yuan JY, Diwan D, Vellimana AK, Osbun JW, Chatterjee AR, Athiraman U, Dhar R, Zipfel GJ. Early brain injury after subarachnoid hemorrhage: incidence and mechanisms. Stroke. 2023;54(5):1426–1440. doi: 10.1161/STROKEAHA.122.040072

14. Korja M, Lehto H, Juvela S, Kaprio J. Incidence of subarachnoid hemorrhage is decreasing together with decreasing smoking rates. Neurology 2016;87(11):1118–1123; doi: 10.1212/WNL.0000000000003091

15. Guo LM, Zhou HY, Xu JW, Wang Y, Qiu YM, Jiang JY. Risk factors related to aneurysmal rebleeding. World Neurosurg. 2011;76(3-4):292–298; discussion 253–254. doi: 10.1016/j.wneu.2011.03.025

16. Weyhenmeyer J, Guandique CF, Leibold A, Lehnert S, Parish J, Han W, Tuchek C, Pandya J, Leipzig T, Payner T, et al. Effects of distance and transport method on intervention and mortality in aneurysmal subarachnoid hemorrhage. J Neurosurg. 2018;128(2):490–498. doi: 10.3171/2016.9.JNS16668

17. Sorteberg A, Romundstad L, Sorteberg W. Timelines and rebleeds in patients admitted into neurosurgical care for aneurysmal subarachnoid haemorrhage. Acta Neurochir (Wien). 2021;163(3):771–781. doi: 10.1007/s00701-020-04673-3

18. Sano H, Satoh A, Murayama Y, Kato Y, Origasa H, Inamasu J, Nouri M, Cherian I, Saito N; members of the 38 registered institutions and WFNS Cerebrovascular Disease & Treatment Committee. Modified World Federation of Neurosurgical Societies subarachnoid hemorrhage grading system. World Neurosurg. 2015;83(5):801–807. doi: 10.1016/j.wneu.2014.12.032

19. Terrett LA, Reszel J, Ameri S, Turgeon AF, McIntyre L, English SW. Elevated blood pressure and culprit aneurysm rebleeding during the unsecured period of aneurysmal subarachnoid hemorrhage: a systematic review. Neurocrit Care. 2024 Oct 14. doi: 10.1007/s12028-024-02138-4

20. Inagawa T, Kamiya K, Ogasawara H, Yano T: Rebleeding of ruptured intracranial aneurysms in the acute stage. Surg Neurol 1987;28(8):93–99, 1987. doi: 10.1016/0090-3019(87)90079-6

21. Doherty RJ, Henry J, Brennan D, Javadpour M. Predictive factors for pre-intervention rebleeding in aneurysmal subarachnoid haemorrhage: a systematic review and meta-analysis. Neurosurg Rev. 2022;46(1):24. doi: 10.1007/s10143-022-01930-0

22. Aladawi M, Elfil M, Ghozy S, Najdawi ZR, Ghaith H, Alzayadneh M, Rabinstein AA, Hawkes MA. The impact of systolic blood pressure reduction on aneurysm re-bleeding in subarachnoid hemorrhage: A systematic review and meta-analysis. J Stroke Cerebrovasc Dis. 2024;33(12):108084. doi: 10.1016/j.jstrokecerebrovasdis.2024.108084

23. Dissanayake AS, Ho KM, Phillips TJ, Honeybul S, Hankey GJ. Pre-treatment re-bleeding following aneurysmal subarachnoid hemorrhage: A systematic review of published prediction models with risk of bias and clinical applicability assessment. J Clin Neurosci. 2024;119:102–111. doi: 10.1016/j.jocn.2023.10.020

24. Kaneko J, Tagami T, Tanaka C, Kuwamoto K, Sato S, Shibata A, Kudo S, Kitahashi A, Kuno M, Yokobori S, et al. Ultra-early induction of general anesthesia for reducing rebleeding rates in patients with aneurysmal subarachnoid hemorrhage. J Stroke Cerebrovasc Dis. 2021;30(8):105926. doi: 10.1016/j.jstrokecerebrovasdis.2021.105926

25. De Marchis GM, Lantigua H, Schmidt JM, Lord AS, Velander AJ, Fernandez A, Falo MC, Agarwal S, Connolly ES Jr, Claassen J, et al. Impact of premorbid hypertension on haemorrhage severity and aneurysm rebleeding risk after subarachnoid haemorrhage. J Neurol Neurosurg Psychiatry. 2014;85(1):56–59. doi: 10.1136/jnnp-2013-305051

26. Haripottawekul A, Persad-Paisley EM, Paracha S, Haque D, Shamshad A, Furie KL, et al. Comparison of the effects of blood pressure parameters on rebleeding and outcomes in unsecured aneurysmal subarachnoid hemorrhage. World Neurosurg. 2024 May;185:e582*–*e590. doi: 10.1016/j.wneu.2024.02.078

27. Kobata H, Ikawa F, Sato A, Kato Y, Sano H. Significance of Pupillary Findings in Decision Making and Outcomes of World Federation of Neurological Societies Grade V Subarachnoid Hemorrhage. Neurosurgery. 2023;93(2):309–319. doi: 10.1227/neu.0000000000002410

28. Kobata H, Sugie A, Tucker A, Sarapuddin G, Kimura H, Takeshita H, Morihara M, Kawakami M. High plasma D-dimer levels correlate with ictal infarction and poor outcomes in spontaneous subarachnoid hemorrhage. World Neurosurg. 2024;190:e809–e822. doi: 10.1016/j.wneu.2024.08.016

29. Kobata H, Sugie A, Yoritsune E, Miyata T, Toho T. Intracranial extravasation of contrast medium during diagnostic CT angiography in the initial evaluation of subarachnoid hemorrhage: report of 16 cases and review of the literature. Springerplus. 2013 Aug 28;2(1):413. doi: 10.1186/2193-1801-2-413

30. Naidech AM, Janjua N, Kreiter KT, Ostapkovich ND, Fitzsimmons BF, Parra A, Commichau C, Connolly ES, Mayer SA. Predictors and impact of aneurysm rebleeding after subarachnoid hemorrhage. Arch Neurol. 2005;62(3):410–416. doi: 10.1001/archneur.62.3.410. PMID: 15767506

31. Tang C, Zhang TS, Zhou LF: Risk factors for rebleeding of aneurysmal subarachnoid hemorrhage: a meta-analysis. PLoS One 9:e99536, 2014 doi: 10.1371/journal.pone.0099536

32. Connolly ES Jr, Rabinstein AA, Carhuapoma JR, Derdeyn CP, Dion J, Higashida RT, Hoh BL, Kirkness CJ, Naidech AM, Ogilvy CS, et al; Council on Cardiovascular Surgery and Anesthesia; Council on Clinical Cardiology. Guidelines for the management of aneurysmal subarachnoid hemorrhage: a guideline for healthcare professionals from the American Heart Association/american Stroke Association. Stroke. 2012;43(6):1711–1737. doi: 10.1161/STR.0b013e3182587839

33. Steiner T, Juvela S, Unterberg A, Jung C, Forsting M, Rinkel G; European Stroke Organization. European Stroke Organization guidelines for the management of intracranial aneurysms and subarachnoid haemorrhage. Cerebrovasc Dis. 2013;35(2):93–112. doi: 10.1159/000346087

34. Hoh BL, Ko NU, Amin-Hanjani S, Chou SH-Y, Cruz-Flores S, Dangayach NS, et al. 2023 Guideline for the management of patients with aneurysmal subarachnoid hemorrhage: A guideline from the American Heart Association/American Stroke Association. Stroke. 2023 Jul;54(7):e314–e370. doi: 10.1161/STR.0000000000000436

35. Treggiari MM, Rabinstein AA, Busl KM, Caylor MM, Citerio G, Deem S, Diringer M, Fox E, Livesay S, Sheth KN, et al. Guidelines for the Neurocritical Care Management of Aneurysmal Subarachnoid Hemorrhage. Neurocrit Care. 2023;39(1):1–28. doi: 10.1007/s12028-023-01713-5

36. Hofmann BB, Donaldson DM, Fischer I, Karadag C, Neyazi M, Piedade GS, Abusabha Y, Muhammad S, Rubbert C, Hänggi D, et al. Blood pressure affects the early CT perfusion imaging in patients with aSAH reflecting early disturbed autoregulation. Neurocrit Care. 2023 Aug;39(1):125–134. doi: 10.1007/s12028-023-01683-8

37. Kobata H, Sugie A, Masubuchi T. Management of poor grade subarachnoid hemorrhage. Unsolved problems in the ultra-acute phase. Surg Cereb Stroke (Jpn). 2007;35(4):300–306, 2007. doi: 10.2335/scs.35.300

38. Yamaguchi S, Izumo T, Sato I, Morofuji Y, Kaminogo M, Anda T, Horie N, Matsuo T; Nagasaki SAH Registry Study. Impact of immediate general anesthesia in the emergency room on prevention of rebleeding after subarachnoid hemorrhage. Acta Neurochir (Wien). 2023;165(10):2855–2864. doi: 10.1007/s00701-023-05705-4

